# Long-Term Duration of Antibody Response to SARS CoV-2 in One of the Largest Slums of Buenos Aires

**DOI:** 10.1101/2021.03.05.21253010

**Authors:** Vanina Pagotto, Lorena Luna, Julieta Salto, Magdalena Wagner Manslau, Silvana Figar, Alicia S. Mistchenko, Georgina Carciofi Boyero, Natacha Weinberger, COVIDAR Group, Ana María Gómez Saldaño, Carla Alpire Alponte, Patricia Auza Alarcón, Ayelén Copa Tarqui, Sheila Cortez, Pamela Gallardo, Janeth Gemio Pinaya, Ángeles Hernandez Navarro, Alejandro Maccio, Paula Mosqueda, Nicole Neme, Bania Quispe, Emilio Ramírez Bernal, Thelma Soria, Angélica Fernández Arce, Andrea Gamarnik, Fernán González Bernaldo de Quirós

**Affiliations:** Non-Sponsored Area, Department of Research, Hospital Italiano de Buenos Aires; Community Health, Ministry of Health of Buenos Aires City, Argentina; Population Health Area, Department of Research Hospital Italiano de Buenos Aires; Scientific Research Commission of the Province of Buenos Aires (CIC); Fundación Instituto Leloir-CONICET-Lemos; Primary Health Care, Ministry of Health of Buenos Aires City; Minister of Health, Buenos Aires City, Argentina

## Abstract

The durability of the antibody response following severe acute respiratory syndrome coronavirus 2 (SARS-CoV-2) infections has not been fully elucidated. We have performed a cross-sectional study in one of the largest slums of Buenos Aires, Barrio Padre Mugica in June 2020, detecting a seroprevalence of 53.4%. To evaluate the persistence of the humoral response against SARS-CoV-2 in this area, we designed a second study assessing only the people that were IgG positive in the first survey. The IgG levels against the full spike (S) protein in 175 individuals that were seropositive, at least 6 months before, were evaluated in a second survey. The positivity rate was 92.0%, 161 from 175 individuals remained IgG positive. We observed a contraction in the overall IgG levels measured by ELISA. The median IgG dropped 62% from June to December 2020. Most of the individuals tested (87%) reported to be asymptomatic or oligo-symptomatic. No difference was found between men and women, but people aged less than 50 showed a lower IgG level in each period compared to older individuals. Our data indicate sustained humoral immunity for at least 6 months in a specific socio-economical setting in a population that was mainly asymptomatic for COVID-19.

## Introduction

The COVID-19 pandemic represents the greatest medical and socio-economic challenge of our time (1). Still there are no effective antiviral drugs to treat COVID-19 cases and the recently approved vaccines are not at reach for most of the world population (2–6). It is crucial to understand how long immunity against SARS-CoV-2 persists in infected individuals for decision-making policies and vaccination planning.

Antibody mediated immunity is thought to protect individuals from SARS-CoV-2 by interfering with viral infection. Most of the neutralizing antibodies are directed against the S protein and more specifically to the receptor binding domain RBD (7). Multiple studies have shown that the majority of SARS-CoV-2-infected individuals produce S- and RBD-specific antibodies during the first 2 weeks of the primary response (8). In addition, monoclonal antibodies against this viral protein neutralize the virus *in vitro* and *in vivo (9–11)*. The accumulated information indicates that circulating neutralizing antibodies likely contribute to protection of reinfection, either by their presence in plasma or rapid expression by memory B cells (9,12). In this regard, it has been recently shown that RBD-specific B cells increase and persist up to 8 months post infection (13,14).

Different studies have shown both sustained and declined humoral responses after 6 months of SARS-CoV-2 infection (13,15,16). Several reports have indicated a contraction phase of IgG levels after 2-3 months of infection, followed by a stabilization at relatively high levels over a 6 month period (15). In contrast, other studies have shown a decay in IgG titers as a function of time, reaching undetectable levels during convalescent phases (10).These observations raise a number of questions. Does the longevity of circulating antibodies against SARS-CoV-2 depend on the population assessed? Is it different among inhabitants of wealthy or vulnerable neighborhoods, or other environments such as health care professionals or nursing homes residents? Does the duration of circulating antibodies depend on the mode of viral spread in specific socio-economical settings? With no clear answers to these questions, there is a need to study factors that affect the spread of the virus and the dynamics of the immune responses in different geographic, demographic, and socioeconomic areas.

Here, we focused on evaluating the durability of antibodies in the Barrio Padre Mugica, one of the most vulnerable and overcrowded slums of Buenos Aires City, with more than 40.000 inhabitants (17). The first case of SARS-CoV-2 infection in this Barrio was reported four days after the national mandatory quarantine on March 19th 2020. In June, after a decay in the number of reported SARS-CoV-2 infections in this Barrio, the first seroprevalence survey was performed, and a second study was set in December to evaluate the duration of IgG among seropositive individuals identified in the first survey.

## Materials and Methods

### Ethics statement

The study was approved by the institutional review board of the “Hospital de Niños, Dr. Ricardo Gutierrez”, Argentina. Informed consent was obtained from every participant. The study protocol was registered in clinicaltrials.gov. NCT04673279

### Population and settings

The study was performed in Barrio Padre Mugica, one of the most overcrowded slums with more than 40000 inhabitants, and almost 1500 people experiencing homelessness. The Barrio is located next to the main passenger transfer center of the City of Buenos Aires and near the neighborhoods named Retiro and Recoleta. It is a heterogeneous slum, with 10 different geographical sectors, each presenting particular cultural and social characteristics. Areas: Bajo Autopista, Comunicaciones, Cristo Obrero, Ferroviario, Güemes, Inmigrantes, Playón Este, Playón Oeste, San Martín and YPF. The cohort included residents above 14 years that were positive IgG against SARS-COV-2 in June 2020.

The cross-sectional survey was carried out between December 3er and December 19th 2020, 8 months after the initial reported COVID-19 cases in Barrio Padre Mugica, and 6 months after the first seroprevalence study (18).

### Data collection and analysis methods

The survey collected information of the following variables: age, biological sex, symptoms two months before the study, living and sharing spaces with confirmed COVID-19 case, number of households in the building and the sector of Barrio Mugica. Blood sample collection and epidemiological data were collected and entered in a secure database. The field data was obtained by community health care workers who lived in Barrio Padre Mugica.

Specific anti SARS-CoV-2 IgG levels were measured from blood samples collected by finger pricking. Samples were taken at the doorstep of each household. IgG antibodies against the trimeric S protein were measured using the ELISA COVIDAR test (8). The COVIDAR IgG test was approved by Argentina’s national drug regulatory agency (ANMAT, National Administration for Drugs, Food and Medical Devices) (26). Performance characteristics of the kit indicate a sensitivity of more than 90% after 21 days and more than 95% after 45 days of symptoms onset. The specificity was set to be 100% (8).

### Sample Size and Statistical analysis

In the first survey 426 seropositive individuals were identified. For the second survey, it was defined to reach out 184 individuals from the original 426, in order to detect at least 30% of antibody persistence (5% precision and 95% confidence). A proportionate stratified random sampling was performed in all ten sectors of the Barrio.

Continuous variables were expressed as the mean (standard deviation [SD]) or median (interquartile range (IQR)) and compared by the t-test or Mann-Whitney U test. Categorical variables were expressed as the frequency (%). IgG levels during the follow-up were compared using the Wilcoxon matched pairs signed ranks test. Significant level was set in 0.05 and statistical analysis was performed with software R version 4.0.2

### Funding source

The National and local Ministries of Health provided medical supplies for the survey and for the virology laboratory to perform the ELISA test for SARS-COV-2 antibodies detection. The COVIDAR group provided the Serokits for sampling and the ELISA COVIDAR IgG kits, supported by FOCEM and Asoc. SAND.

None of the funding sources provided economical support for the data collection, statistical analysis, or were used to write the manuscript, or to submit it for publication.

## Results

The first reported case of SARS CoV-2 infection in the Barrio Padre Mugica was reported on March 22nd. From then, reported cases rapidly increased reaching a peak of over 100 cases a day in May 2020 (Fig. 1). In order to study seroprevalence and antibody persistence, two surveys were performed: the first one when the daily reported cases dropped to under 10 cases a day, on June 20 (18), and the second one on December, 7 months from the peak and 6 months from the first survey (Fig. 1).

**Figure 1.**
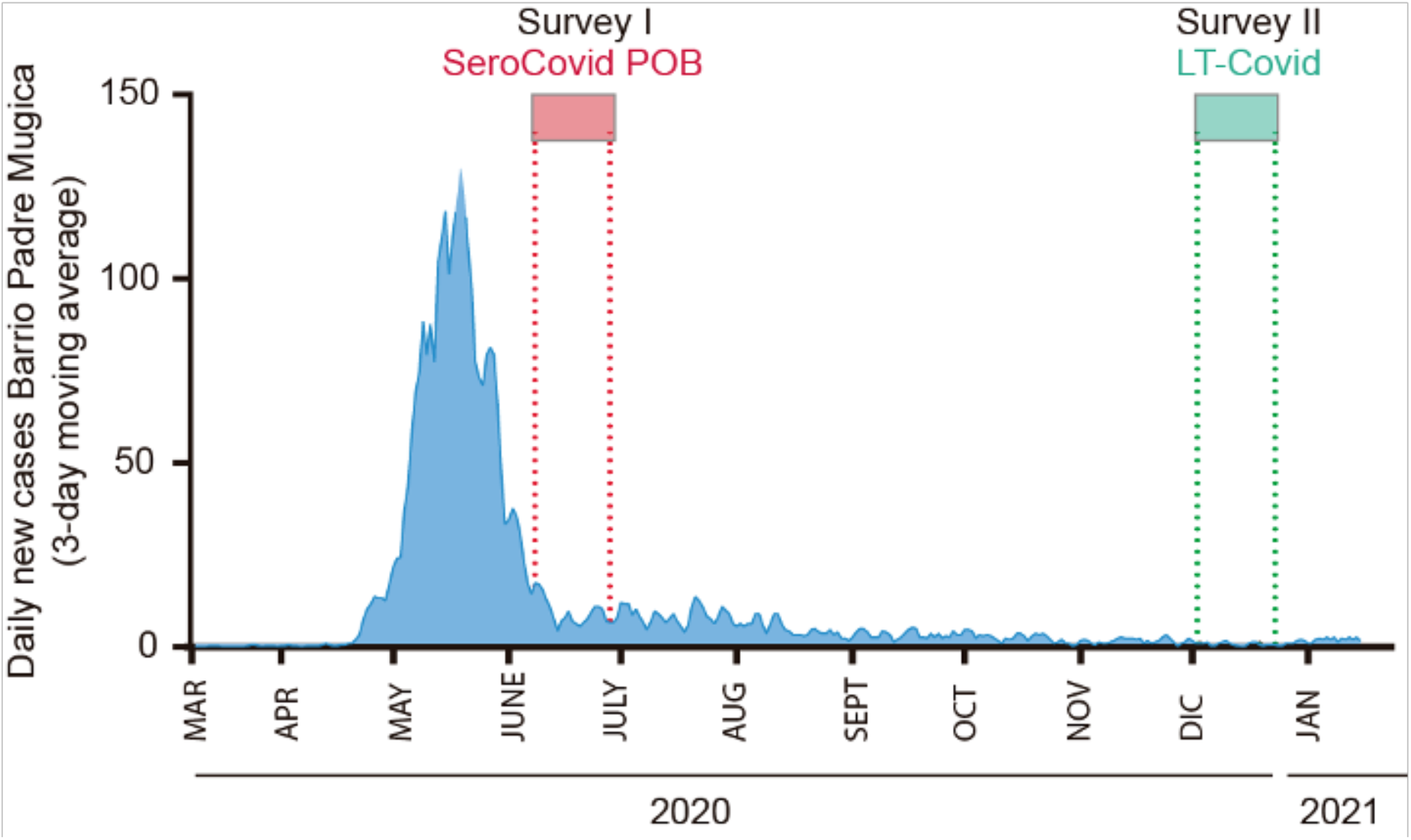
COVID-19 Incident Cases year 2020 at Barrio Padre Mugica, Buenos Aires, Argentina. The two surveys SeroCovid-POB Study (06/2020) and LTCovid Study (12/2020) are indicated on the top.

The survey performed in June 2020 included a cohort of 873 inhabitants of different sectors of the Barrio. ELISA based IgG anti-S of SARS-CoV-2 measurements indicated 421 seropositive cases, resulting in 53.4% seroprevalence (95%IC 52.8% to 54.1%). In December 2020, 220 individuals of the seropositive group were contacted and invited to a second survey, from which 175 agreed to participate, with a response rate of 87%. The field data was obtained by community health care workers who lived in Barrio Padre Mugica. They received training for sampling and collecting survey information. The participatory action research was the groundwork that allowed this study.

The cohort included inhabitants from all 10 areas of the Barrio Padre Mugica, each presenting particular cultural and social characteristics. The amount of individuals selected was based on the population of each area. Regarding the household’s characteristics, the median of people living in the same dwelling was 4 (IQR 2–5) and 55 (31%) shared kitchen and bathroom. Sixty one participant (34.8%) knew a SARS-CoV-2 positive household relative. The persistence of IgG anti-S detected in each area of the Barrio was as follows: Güemes 95% (41 of 43), Playón Oeste 97% (33 of 34), Bajo Playón Este 100% (24 of 24), Ferroviario 75% (12 of 16), San Martín 100% (16 of 16), YPF 57% (8 of 14), Bajo Autopista 100% (10 of 10), Cristo Obrero 100% (11 of 11), Inmigrantes 100% (5 of 5), Comunicaciones 50% (1 of 2) (Fig 2). The study revealed 92% (CI 95% 86.9-95.6) persistence of IgG antibodies anti-SARS. Thus, most of the people that were seropositive in the first survey retained detectable IgGs in the second survey.

**Figure 2.**
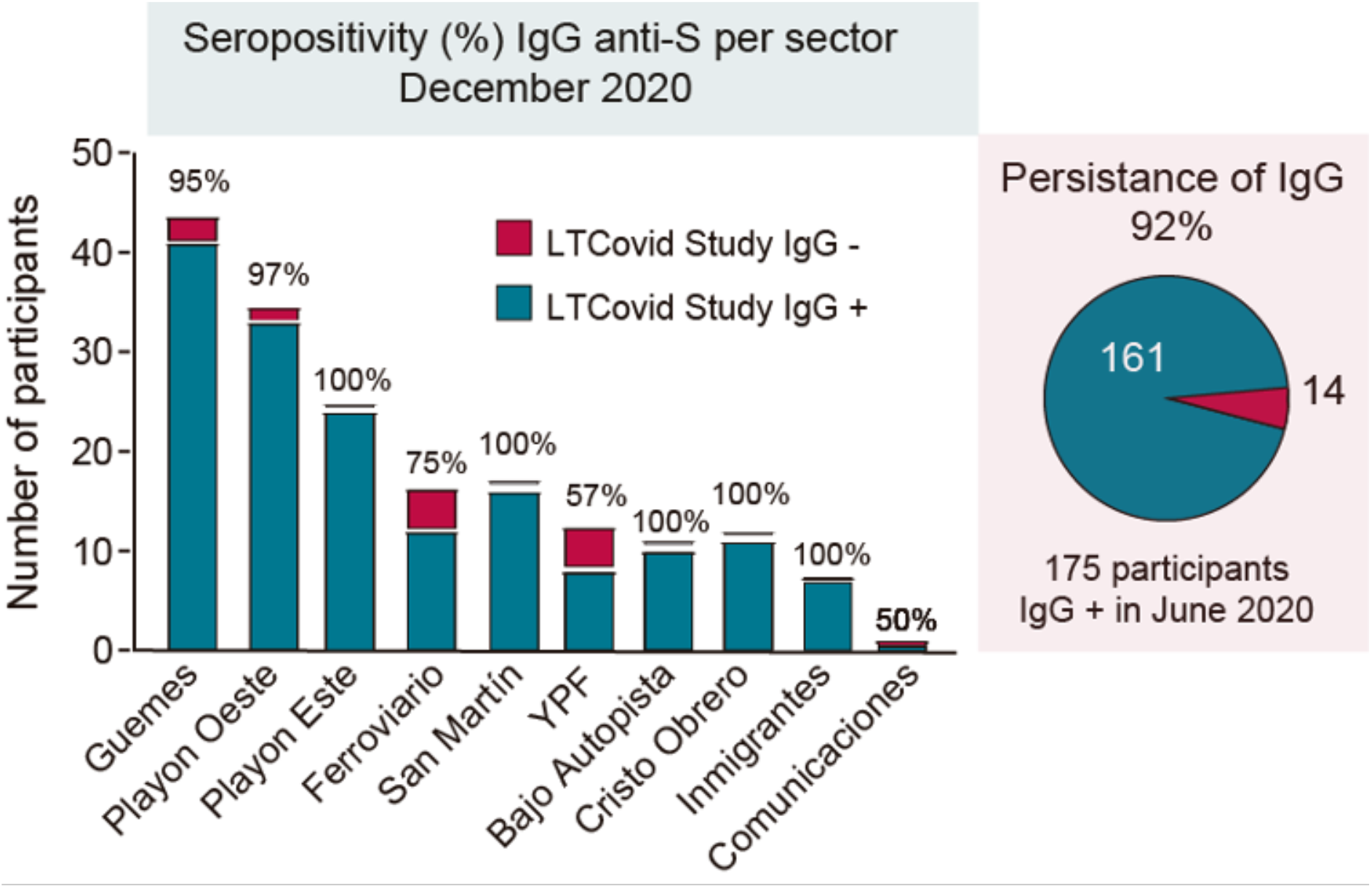
Persistence of IgG anti-S of SARS CoV-2 in each geographical sector of Barrio Padre Mugica in December 2020. Each column represents the total number of individuals assessed and the blue and red colors indicate the IgG positive or IgG negative, respectively. The relative frequency in percentage per sector is indicated on the top. On the right, the overall persistence in the Barrio is shown using the same color code.

It is important to highlight that 140 of the 161 seropositive participants (87%) indicated to be asymptomatic or oligo-symptomatic up to two month before the first survey. This indicates a very high rate of asymptomatic infection in this population. The IgG median level detected in June 2020, measured as optical density OD, was 2.03 (IQR 1.30-3.17). In December 2020 the median OD was 1.26 (IQR 0.69-2.34), indicating a drop of IgG levels of 62% (Fig 3A). Within each study period, no differences in IgG levels were found by sex and presence of symptoms. However, we found that older people (>50 years) displayed greater levels of IgG in both periods compared to people under 50 years (Fig 3B), and both levels declined in the six month period. For >50, IgG levels dropped from median IgG of 3.12 (1.56-3.19) to 1.80 (0.92-2.80) and for <50, from median IgG of 1.93 (1.15-3.00) to 1.18 (0.63-2.04).

**Figure 3.**
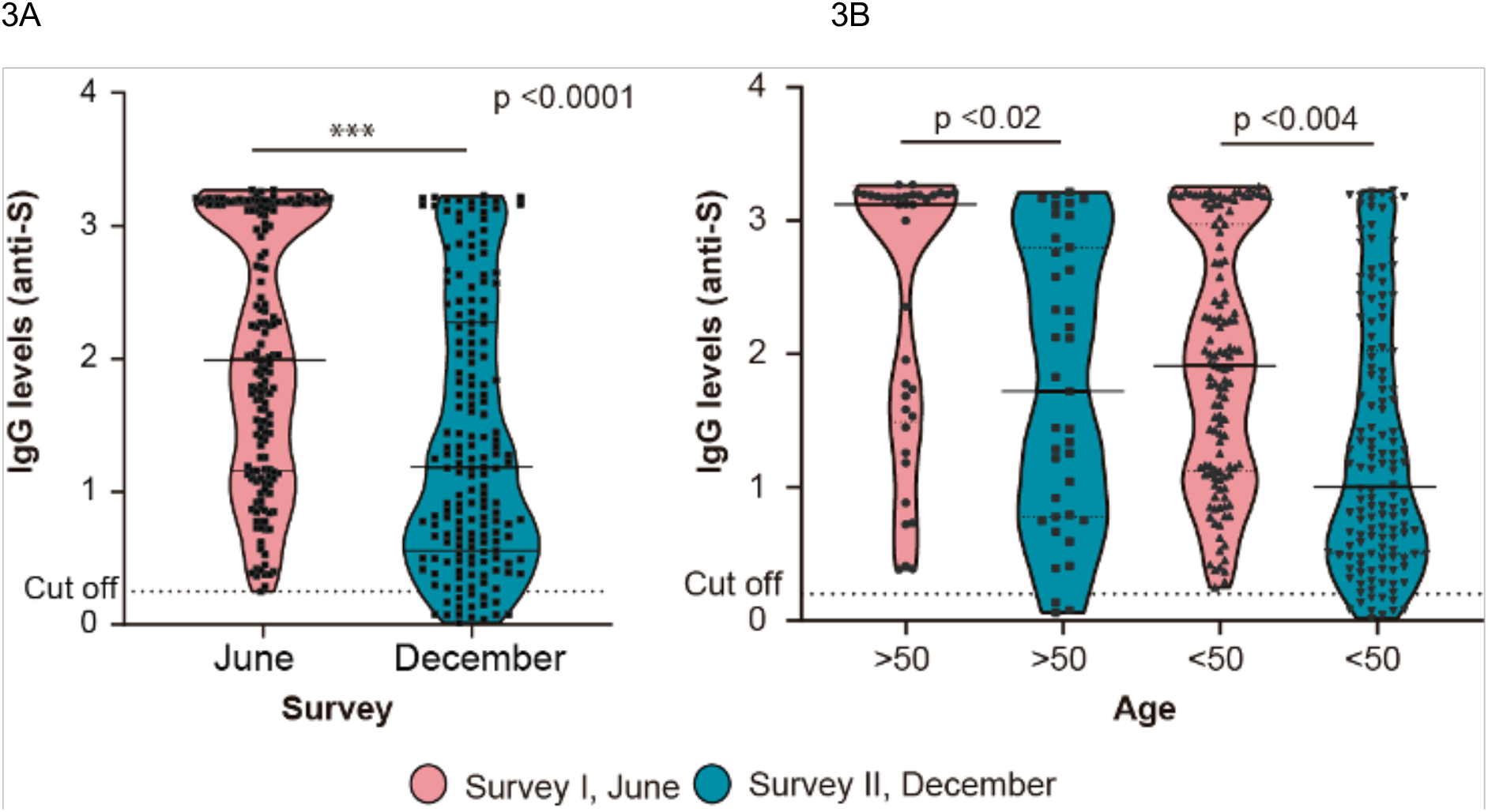
Levels of IgG against Spike of SARS-CoV-2 in SeroCovid-POB study (survey I, June) and in LTCovid Study (Survey II, December). (A) OD levels of IgG measured in serum of the 175 participants in both surveys as indicated. (B) The data shown in A segregated by age as indicated.

In summary, although a significant decay in IgG levels was measured in the observation period, most of the population retained detectable circulating anti-S IgG antibodies.

## Discussion

This is one of the first reports showing IgG anti-SARS persistence in Latin America. We found that 92% of people living in a vulnerable neighborhood in Argentina retained detectable IgG anti-S after at least six month of infection. The persistence of antibodies could be longer (up to 8 months) as the wave peak in that neighborhood occurred before the first cross-sectional study (Fig. 1). The high prevalence of sustained natural immunity found here is of great value for future public health control measures.

This population-based study revealed about 87% of SARS-CoV-2 infected participants without any relevant clinical symptoms. It has been previously reported that asymptomatic individuals showed earlier waning of IgG levels, as compared with symptomatic ones. The immune responses of asymptomatic individuals infected with SARS-CoV-2 have not been well described, mainly because they are more difficult to detect. Here, in a population study of a vulnerable neighborhood, we found a 92% persistence of IgG, which contrasts with that observed in other cohorts. For instance, about 40% of asymptomatic cases, identified as close contacts of qPCR positive cases, were reported to become seronegative in an early convalescent phase (10). These observations suggest that infections in distinct settings may account for different immunological outcomes. We have to bear in mind that different serologic tests used in different studies could yield distinct results. It has been shown with the same samples that anti-S detection displayed higher sensitivity to detect long-lasting IgGs than that with anti-nucleocapsid detection (19).

We found an average contraction of 63% in circulating IgG levels in a six-month period, which is in agreement with previous reports (15). However, it has been shown that a memory phase is triggered when B lymphocytes reactivate antibody production in face of a new infection phase (14,20). In this regard, low or undetectable levels of IgG in convalescent individuals, were found to be followed by a rapid increase in IgG levels after vaccination (21–23). With the recent approval of highly effective vaccines for COVID-19, data on the persistence of immune responses are of central importance. In particular to define whether previously infected people should be vaccinated with full doses and to define priorities in control actions.

The participatory action research process applied here was a cornerstone for gathering the required cohort, and key for accomplishing this study. The “community of inquiry and action” evolved throughout the process, addressing questions that were highly relevant for those who participated in the study. Also, it is important to highlight that co-researchers directly involved in the field work, were community health workers living in the neighborhood (24). This participatory action challenges the notion that the community is a passive object of study, empowering and increasing the knowledge of community members (25).

The current focus for pandemic control is to attain a massive vaccination in a timely fashion. Effective vaccines are being developed and their phase 3 studies are being published. Although their short time efficacy has been already proved for several vaccines, there is not yet guarantee that this protection is long-lasting. Thus, it is essential to evaluate the duration of the immune responses during this world-wide vaccination process and compare it with that in natural infections.

In conclusion, our work provides important data on persistence of anti-SARS-CoV-2 IgG antibodies in a vulnerable neighborhood for at least six months after a wave of infections. The high prevalence of sustained natural immunity found in this study will help to elaborate pandemic control measures.

## Data Availability

not yet decided

